# Cerebrovascular reactivity dispersion as a new biomarker of recent stroke symptomatology in moyamoya

**DOI:** 10.1101/2024.02.27.24303346

**Authors:** Caleb Han, Wesley T. Richerson, Maria Garza, Mark Rodeghier, Murli Mishra, L. Taylor Davis, Matthew Fusco, Rohan Chitale, Shuhei Shiino, Lori C. Jordan, Manus J. Donahue

## Abstract

**Background:** Moyamoya disease (MMD) is a non-atherosclerotic intracranial steno-occlusive condition placing patients at high risk for ischemic stroke. Direct and indirect surgical revascularization can improve blood flow in MMD; however, randomized trials demonstrating efficacy have not been performed and biomarkers of parenchymal hemodynamic impairment are needed to triage patients for interventions and evaluate post-surgical efficacy. We test the hypothesis that hypercapnia-induced maximum cerebrovascular reactivity (CVR_MAX_) and the more novel indicator cerebrovascular reactivity (CVR) response time (CVR_DELAY_), both assessed from time-regression analyses of non-invasive hypercapnic imaging, correlate with recent focal ischemic symptoms.

**Methods:** Hypercapnic reactivity medical resonance imaging (blood oxygenation level-dependent; echo time=35ms; spatial resolution=3.5×3.5×3.5mm) and catheter angiography assessments of cortical reserve capacity and vascular patency, respectively, in MMD participants (n=73) were performed in sequence. Time regression analyses were applied to quantify CVR_MAX_ and CVR_DELAY_. Symptomatology information for each hemisphere (n=109) was categorized into symptomatic (ischemic symptoms within six months) or asymptomatic (no history of ischemic symptoms) and logistic regression analysis assessed the association of CVR metrics with ischemic symptoms after controlling for age and sex.

**Results:** Symptomatic hemispheres displayed lengthened CVR_DELAY_ (p<0.001), which was more discriminatory between hemispheres than CVR_MAX_ (p=0.037). CVR_DELAY_ (p<0.001), but not CVR_MAX_ (p=0.127), was found to be sensitively related to age in asymptomatic tissue (0.33-unit increase/year); age-dependent normative ranges are presented to enable quantitative assessment of patient-specific impairment. Furthermore, the area under the receiver operating characteristic curves shows that CVR_DELAY_ predicts ischemic symptoms (p<0.001), whereas CVR_MAX_ does not (p=0.056).

**Conclusion:** Findings support that CVR metrics are uniquely altered in hemispheres with recent ischemic symptoms, motivating the investigation of CVR as a surrogate of ischemic symptomatology and treatment efficacy.

## Introduction

Moyamoya disease (MMD) is a cerebrovascular condition characterized by the pathogenomic non-atherosclerotic steno-occlusion of the intracranial carotid artery (ICA) and its proximal branches, the corresponding development of lenticulostriate or leptomeningeal collaterals, and high risk of new or recurrent stroke.^1^ Once thought to affect primarily females of East Asian descent, more than 11,000 hospital admissions for patients with a primary diagnosis of MMD are reported annually in the United States and adult MMD admissions have increased more than four-fold in the past 30 years, attributable to increased MMD awareness and possibly prevalence.^2,3^ Adult North American MMD patients are primarily female (3:1 female: male ratio), most often present in their late 30s to early 40s, and are predominately White (48.7%), Black (24.7%), or Hispanic (10.5%).^3^ Ischemic stroke is the initial presentation in 50-60% of these patients,^4^ and despite a lack of randomized trials, treatment most frequently consists of direct or indirect surgical revascularization to improve cerebral perfusion.^5,6^ However, post-operatively a high 16.3% and 23.3% of patients have a new stroke or transient ischemic attack, respectively, and biomarkers that inform stroke risk and personalized management are limited.

Structural medical resonance imaging (MRI) and catheter angiography are frequently used to assess the extent of impairment, the need for direct or indirect surgical revascularization, or the response to treatment.^7^ However, these methods lack information on parenchymal impairment and are increasingly complemented with parenchymal measures of cerebrovascular reserve capacity and microvascular vasodilatory kinetics. Furthermore, while surgical revascularization is the most effective treatment to stabilize or improve cerebrovascular hemodynamics in MMD and secondary moyamoya syndrome (MMS),^8^ treatment response is variable, emphasizing a need for response efficacy biomarkers. Common anatomical imaging methods such as diffusion-weighted imaging (DWI) and FLuid Attenuated Inversion Recovery (FLAIR) MRI demonstrate changes only after irreversible tissue damage, and as such prognostic functional biomarkers are warranted.

Cerebrovascular reactivity (CVR) measures hemodynamic changes in the brain parenchyma in response to a vasoactive stimulus and may provide a functional assessment of how near microvasculature is to exhausting reserve capacity, termed cerebrovascular reserve. While first measured clinically through intravenous infusion of carbonic anhydrase inhibitors (e.g., acetazolamide) and invasive radiological modalities (e.g., ^15^O-positron emission tomography, PET, or single-photon emission computerized tomography, SPECT), owing to procedural risks of these protocols there is an increasing emphasis on assessing CVR using hypercapnic respiratory challenges and blood oxygen level-dependent (BOLD) MRI. Given abilities to modulate the respiratory stimulus quickly and the temporal resolution of BOLD (1-3s), this approach allows for the decomposition of the CVR into the maximum reactivity and reactivity delay time, which may provide unique information on cerebrovascular health. However, for CVR metrics to be accepted as useful biomarkers, the relationship between these measures and ischemic symptomology must be established.

Therefore, the objective of this study is to quantify the sensitivity of hypercapnia-induced CVR metrics with recent ischemic symptoms derived from the same flow territory using novel time-regression analyses.^9,10^ We hypothesize that the cortical cerebrovascular reactivity timing profile (i.e., the delay of response) is more sensitive to acute or early chronic ischemic symptomatology compared to the magnitude of the vasodilatory response itself. Furthermore, this study aims to provide exemplary statistics to enable personalized, diagnostic reactivity assessments.

## Materials and Methods

### Demographic and study design

All components of this study were in compliance with the Declaration of Helsinki of 1975 (and as revised in 1983) and Health Insurance Portability and Accountability Act (HIPPA), were approved by Vanderbilt University Medical Center Institutional Review Board, and followed the Strengthening the Reporting of Observational studies in Epidemiology (STROBE) guidlines.^11^ Participants (n=73) with moyamoya disease or syndrome were identified from the neurology and/or neurological surgery services at Vanderbilt University Medical Center, and all participants provided informed, written consent in compliance with the Vanderbilt University Medical Center Institutional Review Board (IRB Study #160478). Inclusion criteria were a clinical diagnosis of moyamoya vasculopathy based on catheter angiography and neurological assessment.^1^

### Neurological assessment

A board-certified neurologist (experience = 17 years) organized each participant’s symptomatology information for each hemisphere into symptomatic (focal ischemic symptoms within six months of their respective BOLD MRI scan) or asymptomatic (no history of ischemic symptoms) cohorts. Ischemic symptoms included stroke or transient ischemic attack documented in the chart or radiological indicators of focal ischemic injury, like the presence of new infarcts within the last 6 months. Hemispheres that did not meet either criterion (i.e., hemispheres that displayed ischemic symptoms greater than six months ago) were excluded from the study.

### Catheter angiography

Pre-surgical six-vessel digital subtraction angiography was performed on each participant using a Philips Allura Xper biplane neuro X-ray system with the participant in the supine position. The collaborating neurosurgeons performed internal carotid artery injections (6 cc/s, 8 cc injected), external carotid artery injections (3 cc/s, 6 cc injected), and common carotid artery injections (8 cc/s, 12 cc injected) using a nonionic, water-soluble intra-arterial contrast. Digital images were acquired at approximately three frames per second.^12^

### MRI acquisition

To evaluate hemodynamic reactivity, BOLD data were acquired (TE=35 ms; spatial resolution=3.5×3.5×3.5 mm, TR=2000 ms) at 3.0 Tesla (Philips, Philips Healthcare, Best, The Netherlands) using body coil radiofrequency transmission and SENSE phased-array reception.^12^ Given long vasodilatory equilibrium times in many patients with MMD, we followed our previously reported protocol where a paradigm of two blocks of 180s hypercapnia (carbogen gas (5% CO_2_/95% O_2_) delivered at 12L/min through an oxygen face mask) is interleaved with 180s normocapnia (∼21% O_2_ / ∼79% N_2_ delivered at the same flow rate and procedure), with a beginning and ending 90s of normocapnia.^13^ All gases were certified for human consumption and heart rate, blood pressure, arterial oxygen saturation (SaO_2_), and end-tidal carbon dioxide (EtCO_2_) were monitored using an MR-compatible monitoring system (Invivo, Orlando, FL, USA) during MRI acquisition.

### MRI analysis

Functional MRI data were corrected for motion and baseline drift.^13^ BOLD preprocessing included motion correction using Functional Magnetic Resonance Imaging of the Brain (FMRIB) Software Library (FSL) and spatial smoothing with a kernel of full-width-half-maximum = 3mm.^14^ Time regression analysis was performed for quantification of timing uncorrected cerebrovascular reactivity weighted metrics (CVR_RAW_), maximum cerebrovascular reactivity (CVR_MAX_), and reactivity delay time (CVR_DELAY_).^10^ Here, a rectangular regressor (3 min on, 3 min off) to correspond with the timing of the stimulus was progressed in time and voxel-wise z-statistics were calculated for each time progression. The z-statistic map corresponding to the delivery of the respiratory stimulus (e.g., no time progression) was taken as the CVR_RAW_, whereas the maximum z-statistic was recorded as the CVR_MAX,_ and the time at which this maximum z-statistic occurred was recorded as the CVR_DELAY_. In both flow territories, variables preserved for hypothesis testing were the mean z-statistic (e.g., CVR_RAW_) and maximum z-statistic (e.g., CVR_MAX_), each normalized by the change in EtCO_2_ (ΔEtCO_2_).

To obtain cerebral reactivity statistics of the cortical region, masks were derived from the middle cerebral artery (MCA) flow territory and dichotomized along the midline so that the reactivity from the left and right hemispheres could be separately calculated. Furthermore, to ensure the masks only covered healthy tissue, FLAIR and *T*_1_-weighted (T_1_w) images were checked for infarcts, defined as hyperintense on FLAIR and hypointense on T_1_w approaching cerebrospinal fluid (CSF) signal. Tissue meeting infarction criteria was removed from the flow territory masks to ensure that only viable tissue was analyzed.

### Statistical analysis

This study aimed to evaluate the hypothesized correlation between cerebrovascular reactivity parameters (e.g., reduced CVR_RAW_ and CVR_MAX_ and lengthened CVR_DELAY_) and recent ischemic symptomology. Because both hemispheres for some participants and only one hemisphere for others were included in the study, standard errors that account for clustering are calculated for all statistical tests. Demographic variables and reactivity measures are summarized with the mean and standard deviation for continuous variables and accounts and percentages for categorical variables. A logistic regression analysis, with controls for age and sex, was used to assess the association between the CVR metrics and symptomatology. The model used robust standard errors based on the Huber-White sandwich estimator to adjust for clustering.

To compare the ability of each reactivity measure to identify recently symptomatic hemispheres, receiver operating characteristic (ROC) analysis was completed for each, including the ROC curves, and the area under the curves (AUC). Additionally, a linear regression analysis was performed for each CVR metric with age in asymptomatic hemispheres to provide an example of how CVR can be utilized as a diagnostic motivator. The criterion for significance for models was two-sided p<0.05 (p<0.017 after Bonferroni correction for testing three CVR statistical measures).

## Results

Figure 1 displays the schematic of the enrollment process. FLAIR images and the corresponding CVR maps are shown of a participant in their 30s with recent left ischemic symptoms to serve as a visual aid of how the hemispheres are divided and organized into cohorts.

**Figure 1.**
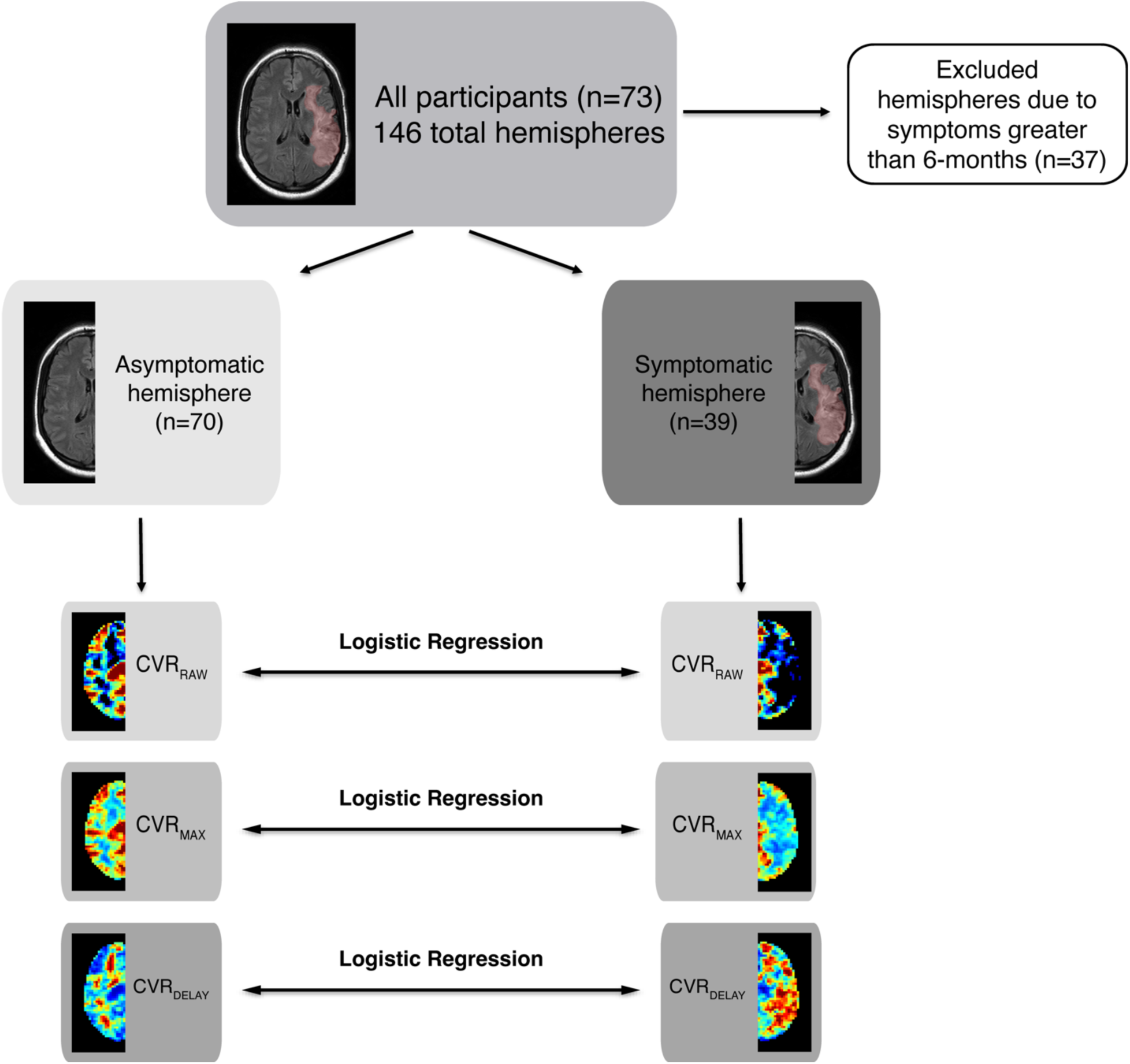
Visual schematic of enrollment and analysis. A schematic diagram of enrollment is shown with fluid-attenuated inversion recovery (FLAIR) and cerebrovascular reactivity (CVR) images of a representative participant in their 30s with recent left hemispheric ischemic symptoms. The large left infarct is manually highlighted in red and the corresponding CVR images show reduced CVR and lengthened CVR_DELAY_ at the location of the infarct. In participants with large infarcts, these infarcts are manually removed from the imaging analysis.

There were 73 participants, 48 female (65.8%) and 25 male (34.2%), with a mean age of 44.4 years (std. dev. 13.8). There were 50 (68.5%) White, 19 (26.0%) Black, and 4 (5.5%) Asian participants. The cohort reflects the known increased prevalence of moyamoya in females and between the ages of 35 and 50 years. Altogether 109 hemispheres were included, 49 (45%) right and 60 (55%) left. Of the 109 hemispheres, 39 hemispheres (35.8%) were symptomatic and 70 (64.2%) were asymptomatic. Table 1 summarizes the demographic and CVR measures in asymptomatic compared to symptomatic hemispheres. There are no differences in sex, age, or race by symptomatology. No difference was observed in the mean ΔEtCO_2_ between the asymptomatic (5.87±2.09 mmHg) and symptomatic hemispheres (6.43±3.28 mmHg) in response to the hypercapnic challenge (p=0.430). The symptomatic cohort had a significantly higher percentage of participants with idiopathic moyamoya compared to that of the asymptomatic cohort (69.2% to 50.0%, respectively, p=0.041). Within the same flow territories, the symptomatic cohort had a significant reduction of 30.7% in cortical CVR_RAW_ compared to the asymptomatic hemispheres (p=0.003). The symptomatic hemispheres had a 14.2% reduction in cortical CVR_MAX_ (p=0.047) compared to the asymptomatic hemispheres, and the cortical CVR_DELAY_ was increased by 27.7% in the symptomatic hemispheres compared to the asymptomatic hemispheres (p<0.001).

**Table 1.** Participant demographics and reactivity measures, stratified by ischemic symptom history (n=73 participants with 109 left and right hemisphere measurements).

| Variable<sup>#</sup> | Asymptomatic | Symptomatic | p-value* |
| --- | --- | --- | --- |
| N (hemispheres) | 70 | 39 | NA |
| N (participants) | 59 | 32 | NA |
| Age (years) | 44.4 ± 15.4 | 42.8 ± 11.3 | 0.581 |
| Sex, female, count (%) | 45 (64.3) | 27 (69.2) | 0.622 |
| Fraction Idiopathic, count (%) | 35 (50.0) | 27 (69.2) | 0.041 |
| Δ EtCO<sub>2</sub> (mmHg) | 5.87 ± 2.09 | 6.43 ± 3.28 | 0.430 |
| **Race, count (%)** |  |  | 0.291 |
| Asian | 6 (8.6%) | 1 (2.7%) |  |
| Black | 16 (22.9%) | 12 (30.8%) |  |
| White | 48 (68.6%) | 26 (66.7%) |  |
| **Reactivity parameters** |  |  |  |
| Normalized Cerebrovascular Reactivity-Weighted (CVR<sub>RAW</sub>) (z-statistic/ΔEtCO<sub>2</sub>) | 0.75 ± 0.41 | 0.52 ± 0.37 | 0.003 |
| Normalized Maximum Cerebrovascular Reactivity (CVR<sub>MAX</sub>) (z-statistic/ΔEtCO<sub>2</sub>) | 1.74 ± 0.66 | 1.50 ± 0.68 | 0.047 |
| Cerebrovascular Reactivity Delay (CVR<sub>DELAY</sub>) (seconds) | 38.50 ± 10.51 | 49.16 ± 15.19 | <0.001 |

\*Chi-square test for counts; two-sample t-test for means. Tests are adjusted for clustering by participant.
<sup>#</sup>Values are mean (standard deviation) unless otherwise noted.

Table 2 reports the results for the logistic regression models predicting hemispheric symptomatology with the cortical reactivity data, with one model for each CVR measure. Increased CVR_RAW_ was associated with a decreased odds for ischemic symptoms (odds ratio=0.15, p=0.007). After correcting for multiple comparisons, CVR_MAX_ was not associated with the ischemic symptoms (odds ratio=0.54, p=0.038). Increasing CVR_DELAY_ was associated with an increased odds for ischemic symptoms (odds ratio=1.09, p< 0.001). Neither sex nor age were associated with ischemic symptoms.

**Table 2.** Summary of logistic regression and receiver operating characteristic (ROC) analysis of the cerebrovascular reactivity (CVR) metrics. This table summarizes the results of the logistic regression analysis of the reactivity statistics comparing the average (i) cerebrovascular reactivity (CVR_RAW_), (ii) maximum cerebrovascular reactivity (CVR_MAX_), and (iii) cerebrovascular reactivity delay (CVR_DELAY_) among participants who have been stratified by their ischemic symptom history. Furthermore, the results from the AUC-ROC analysis are summarized to show the predictive power of each CVR metric on symptomatology. *p<0.01; **p<0.001.

| **Logistic Regression Models** |  |  |  |  |
| --- | --- | --- | --- | --- |
| **CVR Metrics** | **Variable** | **Odds ratio** | **95% confidence interval** | **P-value** |
| Normalized MCA<br>CVR<sub>RAW</sub> | Age | 0.98 | 0.94 – 1.01 | 0.161 |
|  | Sex - female | 1.24 | 0.47 – 3.30 | 0.661 |
|  | MCA CVR<sub>RAW</sub> | 0.15 | 0.04 – 0.59 | 0.007* |
| Normalized MCA<br>CVR<sub>MAX</sub> | Age | 0.99 | 0.96 – 1.02 | 0.390 |
|  | Sex - female | 1.27 | 0.51 – 3.20 | 0.608 |
|  | MCA CVR<sub>MAX</sub> | 0.54 | 0.30 – 0.97 | 0.038 |
| MCA CVR<sub>DELAY</sub> | Age | 0.96 | 0.93 – 1.00 | 0.062 |
|  | Sex - female | 1.18 | 0.39 – 3.53 | 0.766 |
|  | MCA CVR<sub>DELAY</sub> | 1.09 | 1.05 – 1.12 | <0.001** |
| **ROC Analysis** |  |  |  |  |
| **CVR Metrics** | **Area under Curve (AUC)** | **95% confidence interval** | **P Value** |  |
| MCA CVR<sub>RAW</sub> | 0.675 | 0.568 – 0.783 | 0.003* |  |
| MCA CVR<sub>MAX</sub> | 0.611 | 0.494 – 0.727 | 0.056 |  |
| MCA CVR<sub>DELAY</sub> | 0.707 | 0.603 – 0.812 | <0.001** |  |

Table 2 also summarizes the results from the AUC-ROC analysis. Of the two CVR metrics that were significant in the logistic regression models, the AUC for CVR_RAW_ and CVR_DELAY_ is 0.675 (p=0.003) and 0.707 (p<0.001), respectively. CVR_MAX_ has an AUC of 0.611 (p=0.056), not significantly different than the base of 0.50. The ROC curves for the CVR metrics and the reference curve are displayed in Figure 3d.

Figure 2 visually depicts how the CVR metrics are calculated via the time regression analysis applied to a participant in their 40s with recent ischemic symptoms in the right hemisphere. Figure 2b shows the process whereby a regressor corresponding in time to the hypercapnic stimulus is temporally shifted until it is maximally correlated with the BOLD signal time course to quantify CVR_MAX_ and CVR_DELAY_. While Figure 2 displays the time regression analysis for the mean cortical reactivity for the sake of visualization, this temporal correction is performed in each voxel to create the CVR maps.

**Figure 2.**
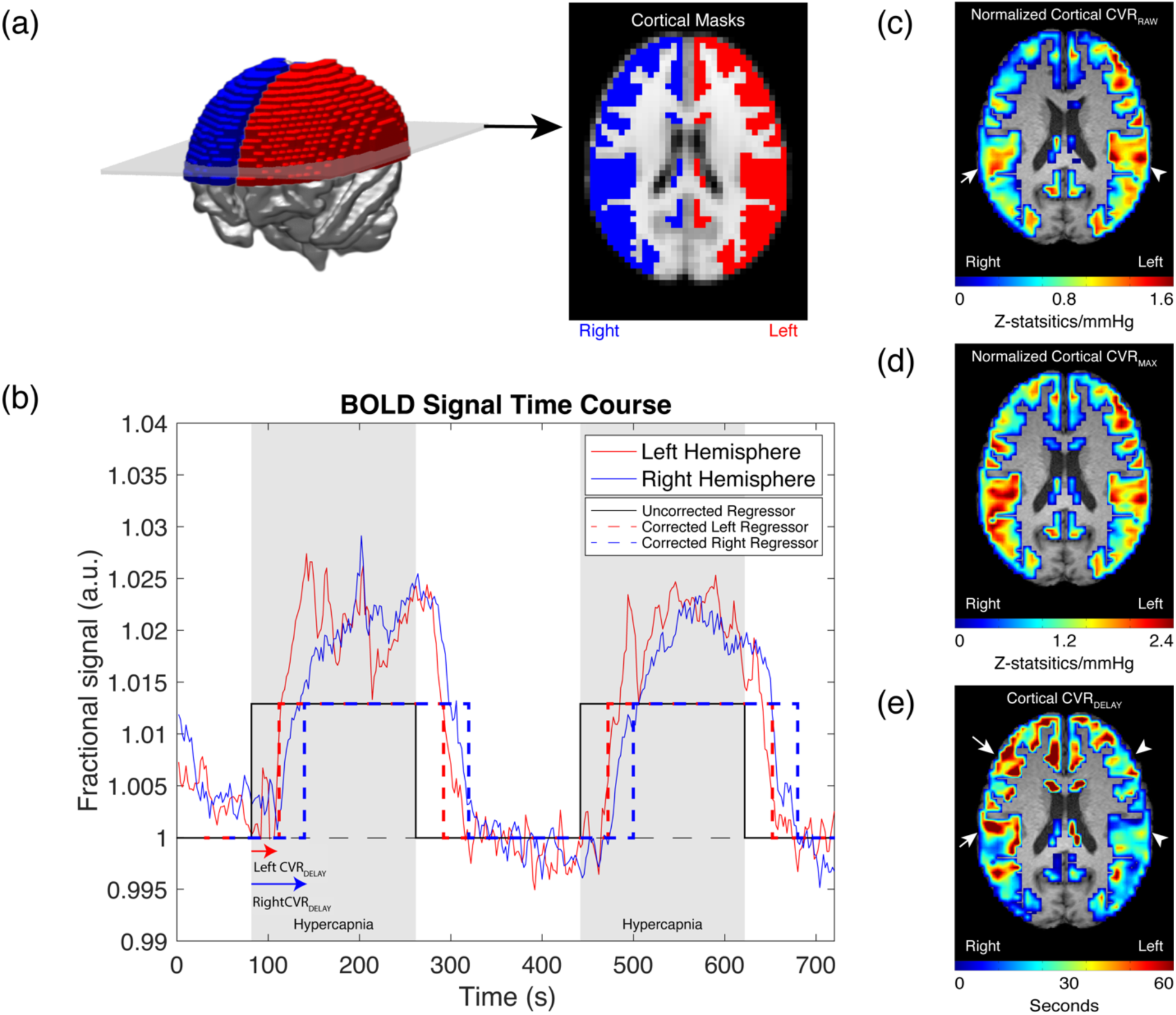
Example of time regression analysis and cerebrovascular reactivity (CVR). (a) A 3-dimensional rendering of the cortical masks is shown with a representative slice that corresponds to primarily anterior (e.g., middle cerebral and anterior cerebral artery) flow territories, where moyamoya vasculopathy is primarily localized. (b) An example of the cortical blood oxygenation level-dependent (BOLD) time course is shown on a representative participant in their 40s with recent ischemic symptoms on the right hemisphere. A regressor (black) reflecting the time of the hypercapnic stimulus is temporally shifted to maximally correlate with each hemisphere’s cortical BOLD time courses (red and blue dotted lines). Both hemispheres display robust signal change, but the right hemisphere shows a lengthened delay in response to the hypercapnic stimulus. (c-e) Examples of the CVR maps at the corresponding level in panel (a) are shown for the same participant. White arrows demarcate reduced CVR_RAW_ and lengthened CVR_DELAY_ in the right hemisphere, arrowheads show a more normal pattern in the left hemisphere.

**Figure 3.**
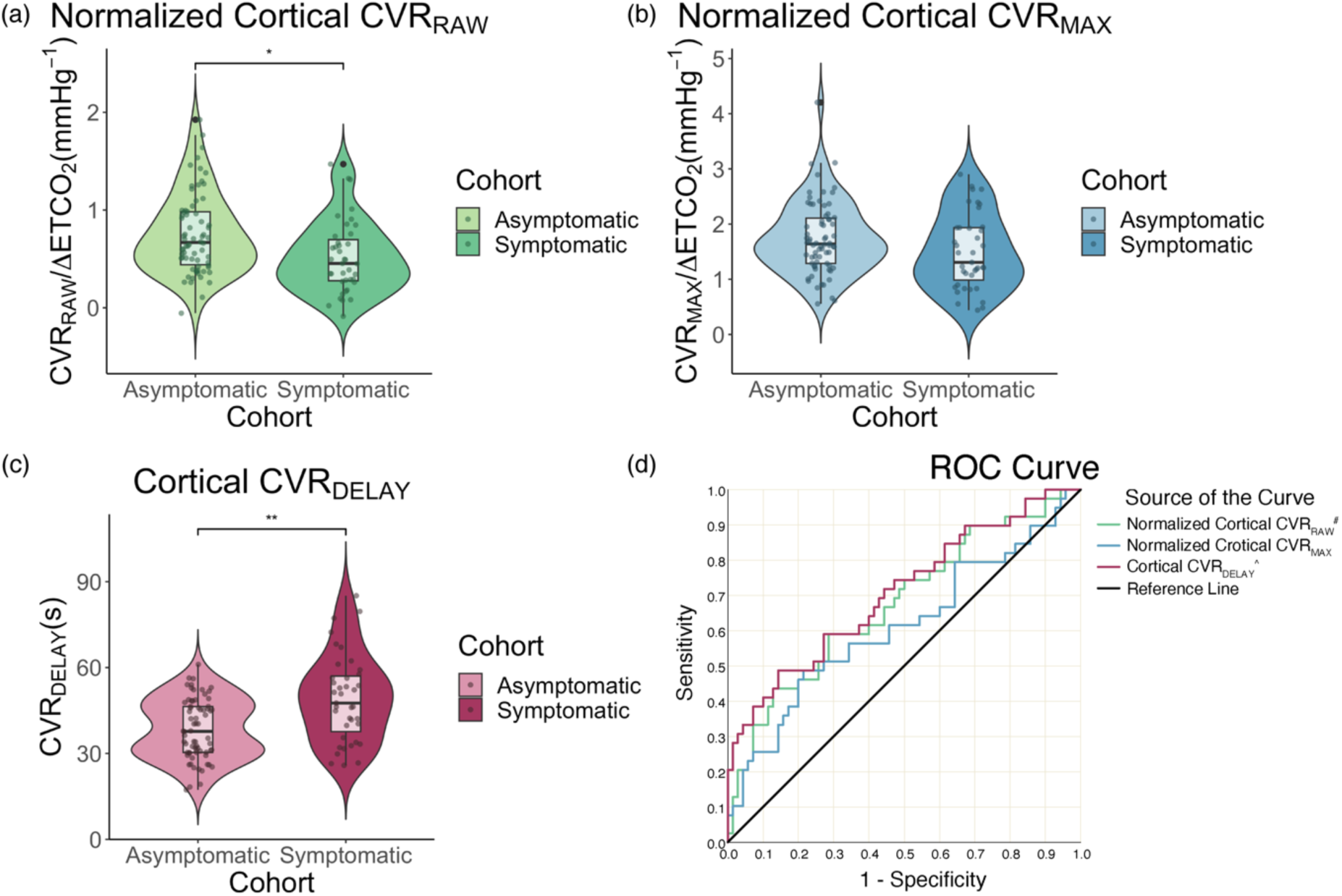
Violin plot comparing the CVR metrics between each cohort. Violin plots compare the cortical cerebrovascular reactivity between participants who have (symptomatic) and have not (asymptomatic) experienced ischemic symptoms in the prior six months. Upon logistic regression after controlling for age and sex, (a) CVR_RAW_ was significantly lower, and (c) the CVR_DELAY_ significantly lengthened in the symptomatic cohort (*p=0.003; **p<0.001). (b) CVR_MAX_ was reduced in symptomatic hemispheres (p=0.037), which did not meet the stated significance criteria after accounting for multiple comparisons. (d) The receiver operating characteristic (ROC) curve for each of the CVR metrics shows statistical evidence that CVR_RAW_ and CVR_DELAY_ have predictive power of symptomatology while CVR_MAX_ does not (^#^p=0.003, ^^^p<0.001).

Figure 3 provides additional information on the data distribution of each cortical CVR metric for the asymptomatic and symptomatic cohort. The violin plots demonstrate the spread of data between cohorts and the group level finding that lower CVR_RAW_, lower CVR_MAX_, and longer CVR_DELAY_ are present in symptomatic vs. asymptomatic parenchyma, which is reported quantitatively in Table 1.

To demonstrate the ability to visualize changes on an individual patient basis, Figure 4 summarizes a case example of a participant in their 50s who had a recent right MCA territory stroke but no history of left hemispheric flow-limiting stenosis or infarct. The angiograms in Figure 4b-c display the severe stenosis of the right MCA, but patent left intracranial vasculature. The CVR masks in Figure 4d demonstrate the hemispheric bias of the corresponding reactivity metrics, with the ischemic and symptomatic hemisphere displaying evidence of impaired CVR (i.e., lower CVR_RAW_, lower CVR_MAX,_ and longer CVR_DELAY_) compared to the unaffected hemisphere.

**Figure 4.**
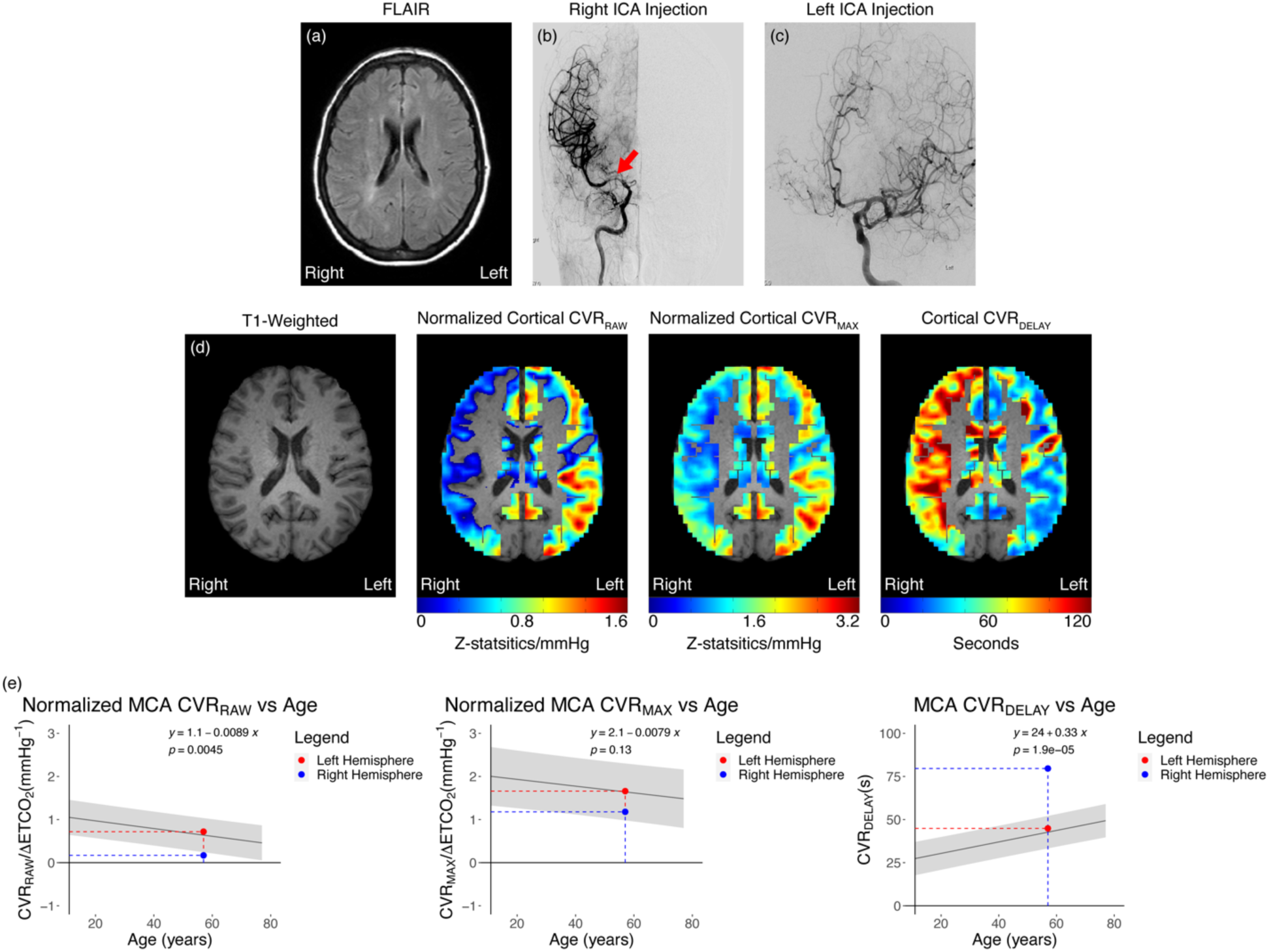
Case example of unilateral stroke. (a) FLAIR image of a participant in their 50s with right hemispheric ischemic symptoms. (b-c) Coronal views of the catheter angiogram of the late arterial phase after bilateral internal carotid (ICA) injections show severe stenosis of the right middle cerebral artery (red arrow). (d) The corresponding *T*_1_-weighted image superimposed with the cortical mask displays reduced reactivity and lengthened reactivity delay in the symptomatic right hemisphere. (e) Plots show age-dependent reactivity regression in asymptomatic hemispheres (n=70) with the average reactivity metrics of the same case example. Asymptomatic (left) hemisphere reactivity metrics fall within the normative range while the asymptomatic hemisphere shows values outside of the normative range.

Figure 4e also displays the cortical CVR metrics of asymptomatic hemispheres as a function of age. The plots show that with increasing age, CVR_RAW_ decreases (p=0.0045), and CVR_DELAY_ increases (p<0.001). While CVR_MAX_ does appear to decrease with age, this trend was not significant (p=0.127). The gray-shaded region shows an interval one standard deviation away from the line of best fit, appropriate for predictions for individual patients.^15^ Furthermore, this panel demonstrates the mean cortical CVR metrics of the case study to the asymptomatic group trend. Here, it can be appreciated that this participant’s asymptomatic (left) hemisphere’s CVR statistics are within the proposed healthy range (shaded area). By contrast, the symptomatic (right) hemisphere CVR_RAW_ falls below the asymptomatic range, and its CVR_DELAY_ exceeds the asymptomatic range. Linear regression coefficients are presented in the figures to facilitate the use of the information for personalized assessment of hemodynamic impairment.

## Discussion

Cerebrovascular reactivity (CVR) with blood oxygenation level-dependent (BOLD) functional MRI was applied in a cohort of 73 participants with moyamoya to investigate hypothesized relationships between CVR response magnitude and timing and ischemic symptomatology. The main conclusion was that CVR metrics are uniquely altered in hemispheres with recent ischemic symptoms, defined here as focal neurological symptoms within six months. Additionally, the delay in the timing of the CVR response was observed to be most closely related to symptoms. Finally, we provide age-dependent exemplars for CVR for quantitative interpretation of reactivity.

The findings of this study should be considered in the context of our methodology. Conventional CVR mapping techniques require physiological manipulation of CO_2_ levels in blood, such as using respiratory stimuli like breath holding,^16^ hyperventilation,^17^ hypercapnia, or alternative pharmacological manipulation through intravenous acetazolamide injection.^18^

Compliance of breath holding and hyperventilation can be variable, and acetazolamide injection does not allow for timing metrics to be quantified; therefore, our method used a respiratory stimulus of hypercapnic hyperoxia. While custom hypercapnic gas mixtures can be difficult to obtain from most gas vendors, the carbogen gas our technique utilizes is readily available and certified for human consumption. Prior work has observed that CVR responses measured with hypercapnic normoxia (e.g., 5% CO2 and balance room air) and carbogen (e.g., 5% CO2 and 95% oxygen) stimuli are correlated across all brain lobes.^13^ However, it should be noted that the balance of oxygen elicits a BOLD response that is unrelated to reactivity, given the fractional decrease in dHb in capillaries and veins that will result from the hyperoxia balance.^19^ It should be noted that the BOLD response to carbogen, while generally correlated with the response to hypercapnic normoxia, is much larger owing to the non-specific nature of the effect of hyperoxia and as such quantitative values should only be compared between identical stimuli.

Prior literature has provided evidence that CVR-weighted MRI can assay hemodynamic patterns in cerebrovascular diseases like moyamoya and other ischemic cerebrovascular conditions.^9,20^ Our study not only confirms this but also provides additional insight into which CVR metric is the most sensitive to recent ischemic symptoms. This extension is fundamental, as utilizing CVR metrics as a surrogate for ischemic symptom risk or treatment response requires knowledge that the metrics correlate with these clinically established indicators of disease severity. Although most of the CVR metrics are altered by symptomology, it is important to note that the CVR_MAX_ was observed to be less significantly related to symptoms and, in fact, did not meet stated significance criteria after correcting for multiple comparisons. This suggests that the time of vascular compliance, rather than the magnitude of response, may be most sensitively related to ischemic symptoms, giving us additional insight into how moyamoya impacts cerebrovascular kinetics. Moyamoya-associated collaterals are often dilated perforator arteries.^21^ This along with our findings related to the CVR_MAX_ suggests that these vessels’ maximum dilation may be less affected in order to compensate for the impaired cerebral blood flow. However, the fact that the CVR timing profile is significantly correlated with ischemic symptoms may indicate that the time for these vessels to reach maximum dilation is more significantly impaired, and less the magnitude of dilation. Mechanisms of this phenomenon could be related to the impairment of the smooth muscle or vascular endothelium in moyamoya leptomeningeal or lenticulostriate collaterals and could be the topic of future work.

As the CVR timing profile is the most sensitive to recent ischemic symptoms, this motivates using this metric as a gauge of treatment efficacy. Prior work has shown that the CVR metrics are improved by surgical revascularization and can indicate good versus poor response to revascularization in adult moyamoya patients, as verified from neoangiogenesis on catheter angiography.^22,23^ Our findings support this notion by showing that CVR metrics are less impaired in asymptomatic hemispheres in a much larger moyamoya cohort (n=73) and also provide evidence that the temporal parameters of the hemodynamic response function are the most sensitive to the presence of recent ischemic symptoms and have the highest predictive power for such symptoms. The exemplar statistics we show in Figure 5 complement recent reports of CVR ranges in healthy control volunteers.^24^ Additionally, the plots confirm the fact that CVR_DELAY_ is the most sensitive to symptomatology, of the metrics measured, as this metric shows much more obvious impairment in the symptomatic hemisphere compared to the other CVR metrics. While hyperintense signals in DWI images do indicate an ischemic stroke, this imaging technique only provides information on how these strokes acutely affect the brain at a tissue level.^25^ Our finding suggests that while CVR_DELAY_ is highly sensitive to symptomatology, like the hyperintensity in DWI, this metric remains sensitive at least within six months after the stroke, whereas the hyperintensity in DWI only remains hyperintense within the first weeks after the stroke.^26^ Furthermore, CVR_DELAY_ provides additional insight into how the blood vessels and the vascular kinetics are affected by these ischemic episodes, which is beyond the scope of what a DWI image can show. Given these reasons and the highly predictive power of CVR_DELAY_ on symptomatology shown by the AUC-ROC analysis, our study shows the clinical relevance of this metric and how it may be the preferred metric for clinicians to assay cerebrovascular impairment.

The study should also be considered in light of several limitations. First, the most common race in both cohorts was Caucasian, which is consistent with North American moyamoya phenotype; however, moyamoya is also common in East Asia.^27^ Given this and a previous study that found differing surgical outcomes in a multiethnic moyamoya cohort,^23,28^ confirmation in different ethnicities may be required when applying our findings to different racial phenotypes of moyamoya. Second, while our analysis did control for demographic characteristics such as age and sex, additional stroke risk factors (hypercholesterolemia, obesity, diabetes, etc.) were not considered. Future studies that model these contributors might provide additional insight. Given that adult moyamoya vasculopathy generally affects individuals in their 30s and 40s, which is prior to the development of significant intracranial atherosclerosis, this is not believed to be a major confound in the present study. Lastly, while this study does find CVR metrics to be related to symptomatology, we were not powered to evaluate if CVR metrics can predict the future occurrence of ischemic symptoms. Prior work from our group has shown the use of CVR as a predictor for ischemic stroke; however, this study has been limited by its small sample size.^29^ Our findings motivate further longitudinal studies that incorporate a larger cohort to confirm the strength of the predictability of CVR on the development of future ischemic symptoms.

In conclusion, we investigate if the reactivity metrics of anterior flow territories that are supplied directly by arteries affected by moyamoya are related to ischemic symptomatology in moyamoya patients. Our observations support that cortical CVR is more impaired in hemispheres with a history of recent ischemic symptoms. More specifically, CVR_DELAY_, or the time it takes for the microvasculature to react maximally to a vasodilatory stimulus, was observed to be the most sensitive to symptomatology, suggesting its potential clinical relevance as a biomarker of stroke risk or treatment response.

## Data Availability

We will make all of the data for this manuscript available upon request for those who have completed the CITI training.

## Acknowledgements

We would like to thank the National Institutes of Neurological Disorders and Stroke for providing funding for this trial.

## Abbreviations

AUC: area under the curve
BOLD: blood oxygen level-dependent
CSF: cerebrospinal fluid
CVR: cerebrovascular reactivity
CVR_RAW_: uncorrected cerebrovascular reactivity weighted metrics
CVR_MAX_: maximum cerebrovascular reactivity
CVR_DELAY_: cerebrovascular reactivity delay time
DWI: diffusion-weighted imaging
EtCO_2_: end-tidal carbon dioxide
FLAIR: FLuid-Attenuated Inversion Recovery
FSL: Functional Magnetic Resonance Imaging of the Brain (FMRIB) Software Library
HIPPA: Health Insurance Portability and Accountability Act
ICA: intracranial carotid artery
IRB: Institutional Review Board
MCA: middle cerebral artery
MMD: moyamoya disease
MMS: moyamoya syndrome
MRI: medical resonance imaging
PET: positron emission tomography
ROC: receiver operating characteristic
SaO_2_: arterial oxygen saturation
SPECT: single position emission computerized tomography
STROBE: Strengthening the Reporting of Observational studies in Epidemiology
TE: echo time
T_1W_: T_1_-weighted
TR: repetition time

## References

1. Scott RM, Smith ER. Moyamoya disease and moyamoya syndrome. N Engl J Med. 2009;360:1226–1237. doi: 10.1056/NEJMra0804622

2. Starke RM, Crowley RW, Maltenfort M, Jabbour PM, Gonzalez LF, Tjoumakaris SI, Randazzo CG, Rosenwasser RH, Dumont AS. Moyamoya disorder in the United States. Neurosurgery. 2012;71:93–99. doi: 10.1227/NEU.0b013e318253ab8e

3. Kainth D, Chaudhry SA, Kainth H, Suri FK, Qureshi AI. Epidemiological and clinical features of moyamoya disease in the USA. Neuroepidemiology. 2013;40:282–287. doi: 10.1159/000345957

4. Guzman R, Lee M, Achrol A, Bell-Stephens T, Kelly M, Do HM, Marks MP, Steinberg GK. Clinical outcome after 450 revascularization procedures for moyamoya disease. Clinical article. Journal of neurosurgery. 2009;111:927–935. doi: 10.3171/2009.4.JNS081649

5. Arias EJ, Derdeyn CP, Dacey RG, Jr., Zipfel GJ. Advances and surgical considerations in the treatment of moyamoya disease. Neurosurgery. 2014;74 Suppl 1:S116–125. doi: 10.1227/NEU.0000000000000229

6. Starke RM, Komotar RJ, Hickman ZL, Paz YE, Pugliese AG, Otten ML, Garrett MC, Elkind MS, Marshall RS, Festa JR, et al. Clinical features, surgical treatment, and long-term outcome in adult patients with moyamoya disease. Clinical article. J Neurosurg. 2009;111:936–942. doi: 10.3171/2009.3.JNS08837

7. Juttukonda MR, Donahue MJ. Neuroimaging of vascular reserve in patients with cerebrovascular diseases. Neuroimage. 2019;187:192–208. doi: 10.1016/j.neuroimage.2017.10.015

8. Zhang X, Xiao W, Zhang Q, Xia D, Gao P, Su J, Yang H, Gao X, Ni W, Lei Y, Gu Y. Progression in Moyamoya Disease: Clinical Features, Neuroimaging Evaluation, and Treatment. Curr Neuropharmacol. 2022;20:292–308. doi: 10.2174/1570159X19666210716114016

9. De Vis JB, Bhogal AA, Hendrikse J, Petersen ET, Siero JCW. Effect sizes of BOLD CVR, resting-state signal fluctuations and time delay measures for the assessment of hemodynamic impairment in carotid occlusion patients. Neuroimage. 2018;179:530–539. doi: 10.1016/j.neuroimage.2018.06.017

10. Donahue MJ, Strother MK, Lindsey KP, Hocke LM, Tong Y, Frederick BD. Time delay processing of hypercapnic fMRI allows quantitative parameterization of cerebrovascular reactivity and blood flow delays. J Cereb Blood Flow Metab. 2016;36:1767–1779. doi: 10.1177/0271678X15608643

11. Cuschieri S. The STROBE guidelines. Saudi J Anaesth. 2019;13:S31-S34. doi: 10.4103/sja.SJA_543_18

12. Donahue MJ, Ayad M, Moore R, van Osch M, Singer R, Clemmons P, Strother M. Relationships between hypercarbic reactivity, cerebral blood flow, and arterial circulation times in patients with moyamoya disease. J Magn Reson Imaging. 2013;38:1129–1139. doi: 10.1002/jmri.24070

13. Donahue MJ, Dethrage LM, Faraco CC, Jordan LC, Clemmons P, Singer R, Mocco J, Shyr Y, Desai A, O’Duffy A, et al. Routine clinical evaluation of cerebrovascular reserve capacity using carbogen in patients with intracranial stenosis. Stroke. 2014;45:2335–2341. doi: 10.1161/STROKEAHA.114.005975

14. Woolrich MW, Jbabdi S, Patenaude B, Chappell M, Makni S, Behrens T, Beckmann C, Jenkinson M, Smith SM. Bayesian analysis of neuroimaging data in FSL. Neuroimage. 2009;45:S173–186. doi: 10.1016/j.neuroimage.2008.10.055

15. Ross SM. Introduction to probability and statistics for engineers and scientists. Sixth Edition. ed. United Kingdom: Elsevier/Academic Press; 2021.

16. Geranmayeh F, Wise RJ, Leech R, Murphy K. Measuring vascular reactivity with breath-holds after stroke: a method to aid interpretation of group-level BOLD signal changes in longitudinal fMRI studies. Hum Brain Mapp. 2015;36:1755–1771. doi: 10.1002/hbm.22735

17. Bright MG, Bulte DP, Jezzard P, Duyn JH. Characterization of regional heterogeneity in cerebrovascular reactivity dynamics using novel hypocapnia task and BOLD fMRI. Neuroimage. 2009;48:166–175. doi: 10.1016/j.neuroimage.2009.05.026

18. Ogasawara K, Ogawa A, Yoshimoto T. Cerebrovascular reactivity to acetazolamide and outcome in patients with symptomatic internal carotid or middle cerebral artery occlusion: a xenon-133 single-photon emission computed tomography study. Stroke. 2002;33:1857–1862. doi: 10.1161/01.str.0000019511.81583.a8

19. Faraco CC, Strother MK, Siero JC, Arteaga DF, Scott AO, Jordan LC, Donahue MJ. The cumulative influence of hyperoxia and hypercapnia on blood oxygenation and R*₂. J Cereb Blood Flow Metab. 2015;35:2032–2042. doi: 10.1038/jcbfm.2015.168

20. Liu P, Liu G, Pinho MC, Lin Z, Thomas BP, Rundle M, Park DC, Huang J, Welch BG, Lu H. Cerebrovascular Reactivity Mapping Using Resting-State BOLD Functional MRI in Healthy Adults and Patients with Moyamoya Disease. Radiology. 2021;299:419–425. doi: 10.1148/radiol.2021203568

21. Kono S, Oka K, Sueishi K. Histopathologic and morphometric studies of leptomeningeal vessels in moyamoya disease. Stroke. 1990;21:1044–1050. doi: 10.1161/01.str.21.7.1044

22. Watchmaker JM, Frederick BD, Fusco MR, Davis LT, Juttukonda MR, Lants SK, Kirshner HS, Donahue MJ. Clinical Use of Cerebrovascular Compliance Imaging to Evaluate Revascularization in Patients With Moyamoya. Neurosurgery. 2019;84:261–271. doi: 10.1093/neuros/nyx635

23. L Waddle S, Garza M, Davis LT, V Chitale R, R Fusco M, A Lee C, Patel NJ, Kang H, Jordan LC, Donahue MJ. Presurgical Magnetic Resonance Imaging Indicators of Revascularization Response in Adults With Moyamoya Vasculopathy. J Magn Reson Imaging. 2022;56:983–994. doi: 10.1002/jmri.28156

24. Sobczyk O, Battisti-Charbonney A, Poublanc J, Crawley AP, Sam K, Fierstra J, Mandell DM, Mikulis DJ, Duffin J, Fisher JA. Assessing cerebrovascular reactivity abnormality by comparison to a reference atlas. J Cereb Blood Flow Metab. 2015;35:213–220. doi: 10.1038/jcbfm.2014.184

25. Fiebach JB, Schellinger PD, Jansen O, Meyer M, Wilde P, Bender J, Schramm P, Juttler E, Oehler J, Hartmann M, et al. CT and diffusion-weighted MR imaging in randomized order: diffusion-weighted imaging results in higher accuracy and lower interrater variability in the diagnosis of hyperacute ischemic stroke. Stroke. 2002;33:2206–2210. doi: 10.1161/01.str.0000026864.20339.cb

26. Albach FN, Brunecker P, Usnich T, Villringer K, Ebinger M, Fiebach JB, Nolte CH. Complete early reversal of diffusion-weighted imaging hyperintensities after ischemic stroke is mainly limited to small embolic lesions. Stroke. 2013;44:1043–1048. doi: 10.1161/STROKEAHA.111.676346

27. Kim JS. Moyamoya Disease: Epidemiology, Clinical Features, and Diagnosis. J Stroke. 2016;18:2-11. doi: 10.5853/jos.2015.01627

28. Feghali J, Xu R, Yang W, Liew J, Tamargo RJ, Marsh EB, Huang J. Differing Surgical Outcomes in a Multiethnic Cohort Suggest Racial Phenotypes in Moyamoya Disease. World Neurosurg. 2019;128:e865–e872. doi: 10.1016/j.wneu.2019.05.019

29. Juttukonda MR, Davis LT, Lants SK, Waddle SL, Lee CA, Patel NJ, Jordan LC, Donahue MJ. A Prospective, Longitudinal Magnetic Resonance Imaging Evaluation of Cerebrovascular Reactivity and Infarct Development in Patients With Intracranial Stenosis. J Magn Reson Imaging. 2021;54:912–922. doi: 10.1002/jmri.27605

